# Changes in internalizing and externalizing problems in Dutch children and adolescents receiving youth care before and during the COVID-19 pandemic

**DOI:** 10.1101/2023.12.11.23299052

**Authors:** Emma M. Broek, Ronald De Meyer, Rachel van der Rijken, Josjan Zijlmans, Hedy A. van Oers, Michiel A.J. Luijten, Hekmat Alrouh, Arne Popma, Meike Bartels, Robert R.J.M. Vermeiren, Tinca J. C. Polderman, Jacintha M. Tieskens

**Affiliations:** LUMC Curium - Child and Adolescent Psychiatry, Leiden University Medical Center, Leiden, The Netherlands; Praktikon, Nijmegen, The Netherlands; Amsterdam UMC location VU University, Child and Adolescent Psychiatry & Psychosocial Care, Amsterdam, The Netherlands; Amsterdam Public Health, Mental health, Amsterdam, The Netherlands; Amsterdam UMC location University of Amsterdam, Emma Children’s Hospital, Child and Adolescent Psychiatry & Psychosocial Care, Amsterdam, The Netherlands; Amsterdam Public Health, Quality of Care, Amsterdam, the Netherlands; Amsterdam Reproduction and Development, Child Development, Amsterdam, The Netherlands; Amsterdam University Medical Center, Vrije Universiteit Amsterdam, Epidemiology and Data Science, Amsterdam, The Netherlands; Amsterdam Public Health, Methodology and Mental Health, Amsterdam, The Netherlands; Vrije Universiteit Amsterdam, Department of Biological Psychology, The Netherlands; Levvel, Academic Center for Child and Adolescent Psychiatry, Amsterdam, The Netherlands; Youz, Parnassia Psychiatric Institute, The Hague, The Netherlands; Karakter Child and Adolescent Psychiatry University Centre, Nijmegen, The Netherlands; University of Groningen, University Medical Center Groningen, Department of Child and Adolescent Psychiatry & Accare Child Study Center, Groningen, The Netherlands

**Author notes:** CORRESPONDENCE TO: Emma M. Broek (MSc), LUMC Curium, Endegeesterstraatweg 27, 2342 AK Oegstgeest, the Netherlands, 071-5159600. These two authors contributed equally to this work (Tinca J. C. Polderman, Jacintha M. Tieskens).

**Keywords:** COVID-19, child and adolescent mental health, youth care, longitudinal study

## Abstract

**Background:** The COVID-19 pandemic had serious effects on the mental health of children and adolescents. However, it is unclear how the pandemic may have affected treatment effects and outcomes in youth care. We investigated if treatment effects and externalizing and internalizing problems of children and adolescents receiving youth care were affected by the COVID-19 pandemic.

**Methods:** We used data from children and adolescents in youth care (*N* = 1,090, *M_age_* = 12.85 (*SD* = 2.83; range = 8-18 years)). Internalizing and externalizing problems were assessed at the start and end of treatment using the Child Behavior Checklist. We inspected change in internalizing and externalizing problems and clinical status at the end of treatment to investigate treatment effects, and the level of problems at the start and end of treatment. Outcomes were compared between three groups: children treated entirely before the COVID-19 pandemic, children who experienced the transition into COVID-19 measures during treatment, and children treated entirely during the pandemic.

**Results:** We did not find evidence that the pandemic affected treatment effectiveness. However, fewer children who were treated during the pandemic recovered from externalizing problems compared to children treated before the pandemic. Children who received treatment entirely during the pandemic also showed more internalizing and externalizing problems at both the start and end of their treatment, and children who experienced the transition into the pandemic showed elevated externalizing problems at both timepoints.

**Conclusions:** Although the change in internalizing and externalizing problems from start to end of treatment was not affected by the pandemic, our findings that children are entering and leaving care with more problems suggest that child mental health has deteriorated since the pandemic.

## Introduction

The COVID-19 pandemic and its restrictive measures severely disrupted people’s lives, particularly those of young people (Branquinho, Santos, Noronha, Ramiro, & de Matos, 2022; Bruinen de Bruin et al., 2020; Caroppo et al., 2021; Khanna, Cicinelli, Gilbert, Honavar, & Murthy, 2020). In the Netherlands, children and adolescents (hereafter referred to as children) experienced restrictions such as school and childcare facility closures, lockdowns, curfews, social distancing, and quarantine (RIVM, 2023a, 2023b). Restrictions like social distancing and lockdowns also disrupted treatment provision and delivery for children receiving mental health care. As a response to these restrictions the field of mental healthcare experienced a rapid transition to telehealth (Folk et al., 2022; Palinkas et al., 2021; Revet et al., 2023) and mental healthcare workers reported increased psychological problems themselves (Alexiou et al., 2021; Crocker et al., 2023; van Doesum et al., 2023). Mental health professionals also reported overall poorer working conditions as a result of the pandemic (Alexiou et al., 2021; Crocker et al., 2023; Palinkas et al., 2021; Revet et al., 2023; van Doesum et al., 2023). These factors may have resulted in poorer treatment outcomes during the COVID-19 pandemic.

Previous findings have indeed shown that children with mental health problems reported deteriorating mental health outcomes over the course of the pandemic (Kauhanen et al., 2022; Panchal et al., 2021; Samji et al., 2022; Singh et al., 2020). Additionally, both cross-sectional and prospective studies with measurements from both before and during the pandemic have indicated that children experiencing psychological or psychiatric problems reported worse mental health during the COVID-19 pandemic compared to before the pandemic. This includes both internalizing and externalizing problems (Adegboye et al., 2021; Cost et al., 2022; Lopez-Serrano et al., 2021; J. Zijlmans et al., 2021; Josjan Zijlmans et al., 2023). These studies seem to suggest poorer mental health outcomes in these vulnerable children during the COVID-19 pandemic.

However, none of these studies included longitudinal data obtained in separate groups of children who were treated before and during the COVID-19 pandemic. It is therefore difficult to conclude whether children entered care with elevated problem severity during the pandemic compared to before the pandemic, or whether treatment effects or outcomes were affected by the pandemic resulting in poorer mental health at the end of treatment. To do so, data from children treated at different moments during the pandemic is needed, both before and after treatment.

This approach has been taken in a recent study in adults by de Beurs et al. (2022), in which reduction of mental health problems were compared between three groups: adults who received treatment entirely before the COVID-19 pandemic, adults who received treatment partially during the COVID-19 pandemic, and adults who received treatment entirely during the COVID-19 pandemic. Results indicated no diminished effectiveness of treatments during the COVID-19 pandemic, as the reduction in symptoms over time between these three groups was found to be the same (de Beurs et al., 2022).

In this study we used a similar approach as de Beurs et al. (2022) to investigate whether the mental health and treatment of children receiving youth care in the Netherlands was similarly (un)affected during the COVID-19 pandemic. We employed a longitudinal research design in which we compared internalizing and externalizing problems in three groups of children in youth care settings in the Netherlands: 1) Children who received treatment entirely before the COVID-19 pandemic, 2) children who started their treatment before COVID-19 measures were implemented and completed treatment while the pandemic was still in effect, and 3) children who started and completed treatment entirely during the COVID-19 pandemic. We firstly inspected whether treatment effectiveness was affected by the COVID-19 pandemic by testing whether a) the change in symptoms over time and b) the clinical status at the end of treatment differed between these groups. Next, we tested whether children entered care with more problems during the COVID-19 pandemic compared to before the pandemic, and finally we tested whether children who were treated during the COVID-19 pandemic left care with more problems than peers treated before the pandemic.

## Methods

### Participants

We used data from children who were treated in Dutch youth care organizations that participate in the Learning Database Youth (SEJN, 2020). These organizations collect and bring together data on the mental health of children receiving youth care. In this study, children between the ages of 8 and 18 years who received a form of outpatient youth care between 2013 and 2022 were included. Data of 19 youth care institutions were used. These centers are situated in northern (45.9%), eastern (34.1%), southern (3.2%), and western (16.8%) parts of the Netherlands. A total of *N* = 1,090 caregivers of individual children participated in our study, with 85.50% of the informants being female. In our sample 38.53% of the children were female, and 61.47% were male. The mean age of the children was 12.85 (*SD* = 2.83; range = 8-18) years. The average treatment duration was 241.2 (*SD* = 114.0) days. See Table 1 for further characteristics of the study population and the three groups.

**Table 1.** Descriptive results of the three COVID groups.

|  | COVID group |  |  |  | *p* | *Post-hoc* |
| --- | --- | --- | --- | --- | --- | --- |
|  | Before | Transition into | During pandemic |  |  |  |
|  | pandemic (1) | pandemic (2) | (3) |  |  |  |
| Initial sample size (*N*) | 668 | 94 | 328 |  |  |  |
|  | *M* (*SD*) | *M* (*SD*) | *M* (*SD*) | *F*(*df*<sub>within</sub>)<sup>C</sup> | *p* | *Post-hoc* |
| Age at start of treatment | 12.84 (2.88) | 12.06 (2.80) | 13.12 (2.69) | 5.13 (1087) | .006 | 1>2<sup>A</sup>; 1=3; 2<3<sup>A</sup> |
| Treatment duration (days) | 229.5 (101.5) | 348.9 (179.1) | 233.9 (97.5) | 50.22 (1087) | <.001 | 1<2<sup>B</sup>; 1=3; 2>3<sup>B</sup> |
| | *N* (%) | *N* (%) | *N* (%) | $\chi^2$(*df*) | *p* | |
| Sex (female) | 264 (39.52) | 31 (32.98) | 125 (38.1) | 1.52 (2) | .467 |  |
| Sex informant (female) | 577 (86.38) | 77 (81.91) | 278 (84.76) | 1.54 (2) | .464 |  |

Note: <sup>A</sup> $p < .05$ ; <sup>B</sup> $p < .001$ ; <sup>C</sup> $df_{between} = 2$ for all *F*-tests;

### Design and procedure

We employed a longitudinal observational design. Data collection was part of the regular treatment in youth care. Children and their families received outpatient care for a range of problems, such as mental, pedagogical, or educational problems. Caregivers were asked to fill out questionnaires about their children at the start and end of treatment. We exclusively utilized measurements completed by the same informant at both the start and end of treatment and ensured that they were assessed within three months of the treatment’s start- or end date. We used mother ratings (∼82%), or father ratings if these were not available (∼14%). If both mother and father ratings were not available, we used reports from other caregivers, including foster parents (∼2.7%), adoptive parents (∼0.5%), stepparents (∼0.5%), and grandparents (∼0.3%).

### Subgroups used in the analyses

We compared mental health problems between three groups of children in youth care with different treatment times relative to the pandemic in the Netherlands. The approach is similar to the one used by de Beurs et al. (2022). The first group (“Before pandemic”) consisted of children who started and ended treatment prior to March 16, 2020, when COVID-19 restrictions were first implemented in the Netherlands (RIVM, 2023a). The second group (“Transition into pandemic”) consisted of children who started treatment prior to March 16, 2020, and completed treatment after March 16, 2020. Finally, the third group (“During pandemic”) consisted of children who started their treatment after March 16, 2020, and ended their treatment before December 31, 2022. Data inclusion for all three groups was restricted to children with a treatment duration longer than 40 days (short-term treatment) to ensure measurable symptom change. In addition, treatments lasting longer than 1,020 days were excluded, as this was the maximum number of treatment days possible in the “During pandemic” group. In addition, we excluded children who started treatment after the end of COVID-19 pandemic measures in the Netherlands (23-03-2022; RIVM (2023c)). See Figure 1 for a visual overview of these groups.

**Figure 1.**
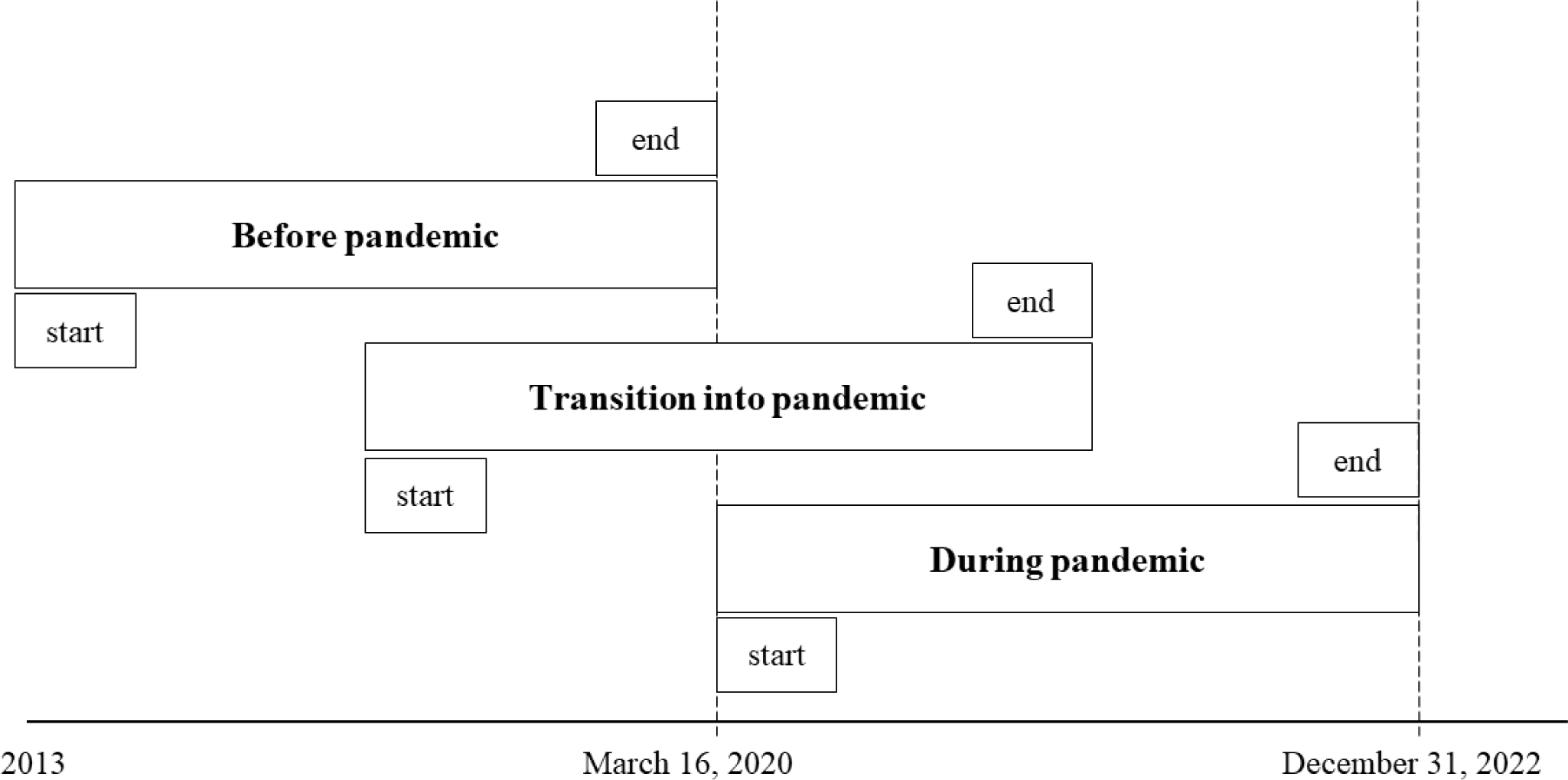
Graphical representation of groups used in statistical analysis based on start and end of treatment relative to March 16, 2020 (i.e. relative to the COVID-19 pandemic).

### Measures

#### Internalizing and externalizing problems

The internalizing and externalizing problem scales of the Child Behavior Check List (CBCL; (Achenbach & Rescorla, 2007) were filled out by caregivers. Items on the CBCL are rated on a three-point Likert scale reflecting how much a statement applies to a child (0 = “not true”; 1 = “somewhat true”; 2 = “very true”). Missing items were scored as 0. To ensure validity of the questionnaires, children with more than 20 missing items of the 120 total problem items were excluded.

The internalizing problems scale consists of 13 items about being anxious/depressed (e.g., “Cries a lot”), 8 items about being withdrawn/depressed (e.g., “There is very little he/she enjoys”), and 11 items about somatic complaints (e.g., “Overtired without good reason”). The total internalizing score was calculated by transforming the total sum score of the 32 items into a *T* score ranging from 33 to 100 (Achenbach & Rescorla, 2007).

The externalizing problems scale consists of 17 items about rule-breaking behavior (e.g., “Drinks alcohol without parents’ approval”) and 18 items about aggressive behavior (e.g., “Destroys his/her own things”). Total externalizing score was calculated by transforming the total sum score of the 35 items into a *T* score ranging from 33 to 100 (Achenbach & Rescorla, 2007).

#### Clinical Status

Two indicators were used to operationalize clinical status at the end of treatment: The Reliable Change Index (RCI), a statistic that determines the magnitude of change of two repeated measures required to be considered statistically reliable (Jacobson & Truax, 1991), and Clinical Significance (CS), the cut-off score to indicate elevated problem scores (Achenbach & Rescorla, 2001).

We considered the reliable change significant when the RCI > 1.65, which corresponds to p < .05. In addition, to decide CS a cut-off score of *T* < 60 was used to categorize a client as functional or *T* ≥ 60 as dysfunctional at the end of treatment. When RCI and CS are combined clients can be categorized into four levels of outcome (Veerman & De Meyer, 2015): 1) Recovered (RCI > 1.65 and CS < 60), 2) improved (RCI > 1.65 and CS ≥ 60), 3) no change (no significant RCI), and 4) deteriorated (RCI < −1.65).

#### Control variables

Age of the child at the start of treatment was measured in years. Sex of both the child and informant was coded as male or female. When questionnaires were filled in by both caregivers, sex was treated as female. Treatment duration was calculated in days by taking the difference between the recorded end and start date of treatment.

### Statistical analysis

To test possible differences in control variables between the three groups, we performed several descriptive analyses. One-way ANOVAs (for child’s age at start of treatment and treatment duration) and χ^2^ tests of independence (for child’s and informant’s sex) were performed to test possible differences between the groups. In addition, correlations between study (internalizing and externalizing problems at beginning and end of treatment, and clinical status at end of treatment) and control (child’s age at start of treatment, treatment duration, sex of both the child and informant) variables were inspected.

To test whether treatment effectiveness was affected by the COVID-19 pandemic we performed two repeated measures ANCOVAs and two χ^2^ tests of independence. The repeated measures ANCOVAs tested whether the change in internalizing and externalizing problems was similar between the three groups. Internalizing and externalizing problems at the start and end of treatment were included as within-subject, time-varying dependent variables and COVID-19 group was included as between-subject independent factor. Age of the child, sex of both the child and informant, and treatment duration were included as covariates. By inspecting the interaction effect between the time-varying factor (i.e., the change in internalizing and externalizing problems over time) and the between-subject factor (i.e., the three groups) we can determine whether the change in problem severity is different between the three groups. χ^2^ tests of independence were used to assess between-group differences in clinical status (recovered, improved, no change, deteriorated) at the end of treatment. Two χ^2^ tests of independence were conducted to test between-group differences in clinical status for internalizing and externalizing problems separately. Post-hoc testing was used to test specific subgroup differences in clinical status proportions if the main χ^2^ test of independence was found to be significant.

To test whether children entered care with elevated problems during the COVID-19 pandemic compared to before the pandemic two ANCOVAs were used to assess between-group differences in internalizing and externalizing problems at the beginning of treatment. Internalizing or externalizing problems at the start of treatment were included as dependent factors, and group as independent factor. Age of the child, sex of both the child and informant, and treatment duration were included as covariates. Post-hoc *t*-tests were used to test between-group differences. To test whether children who were treated during the COVID-19 pandemic left care with more problems compared to peers treated before the pandemic two ANCOVAs were used to assess between-group differences in internalizing and externalizing problems at the end of treatment. Internalizing or externalizing problems at the end of treatment were included as dependent factors, and group as independent factor. Age of the child, sex of both the child and informant, and treatment duration were included as covariates. Post-hoc *t*-tests were used to test between-group differences.

## Results

### Descriptive results

Table 1 presents demographic characteristics of the three groups. The “Before pandemic” group was the largest group (*N* = 668), followed by the “During pandemic” group (*N* = 328), and “Transition into pandemic” group (*N* = 94). Children in the “Transition into pandemic” group were significantly younger (*p* = .006, *M_age_* = 12.06 (*SD* = 2.80)) and had significantly longer treatment duration (*p* < .001, *M_treatment_* = 348.9 days (*SD* = 179.1)) than children in both the “Before pandemic” group (*M_age_* = 12.84 (*SD* = 2.88); *M_treatment_* = 229.5 days (SD = 101.5)) and “During pandemic” group (*M_age_* = 13.12 (*SD* = 2.69); *M_treatment_* = 233.9 days (*SD* = 97.5)). The distribution of sex in participants or informants did not differ between the three groups. In Supplementary Table 1 correlations between all study and control variables are depicted.

### Internalizing problems

In all groups, there was a significant decrease in internalizing problems (*F*(1,1083) = 8.55, *p* = .004, partial η^2^ < .01). The change in internalizing problems from start to end of treatment did not differ between the three groups (*F*(2,1083) = 0.44, *p* = .64, partial η^2^ < .001). See Figure 2A for a graphical representation of these results. Clinical status based on internalizing problems did not differ between the three pandemic groups at the end of treatment (χ^2^(6) = 7.59, *p* = .270, see Table 2). Internalizing problems at the beginning (*F*(2,1083) = 5.57 *p* = .004, η^2^ = .01) and at the end (*F*(2,1083) = 6.84, *p* = .001, η^2^ = .01) of treatment were significantly different between the three groups. Post-hoc *t*-tests revealed that internalizing problems in the “During pandemic” group were significantly higher than in the “Before pandemic” group at both the beginning and end of treatment. No other between-group differences were found at either time point (see Table 3).

**Figure 2.**
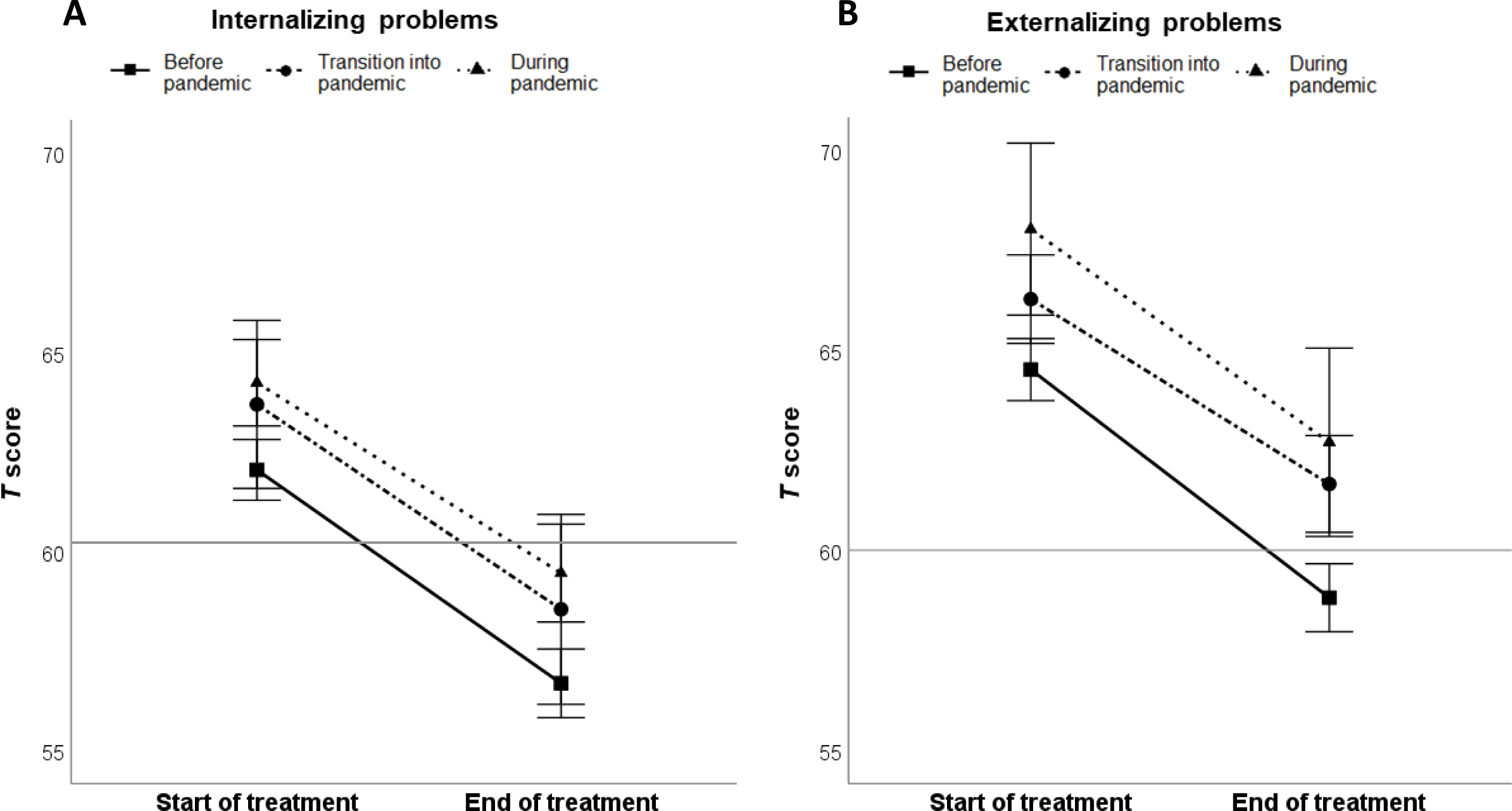
Change over time of (A) internalizing problems and (B) externalizing problems (measured with the CBCL) for the three groups relative to the COVID-19 pandemic. The horizontal line at T score = 60 shows the cutoff for internalizing and externalizing problems at clinical levels **Note:** Internalizing and externalizing problems at start and end of treatment are reported corrected for child’s and informant’s sex, child’s age at start of treatment, and treatment duration

**Table 2.**
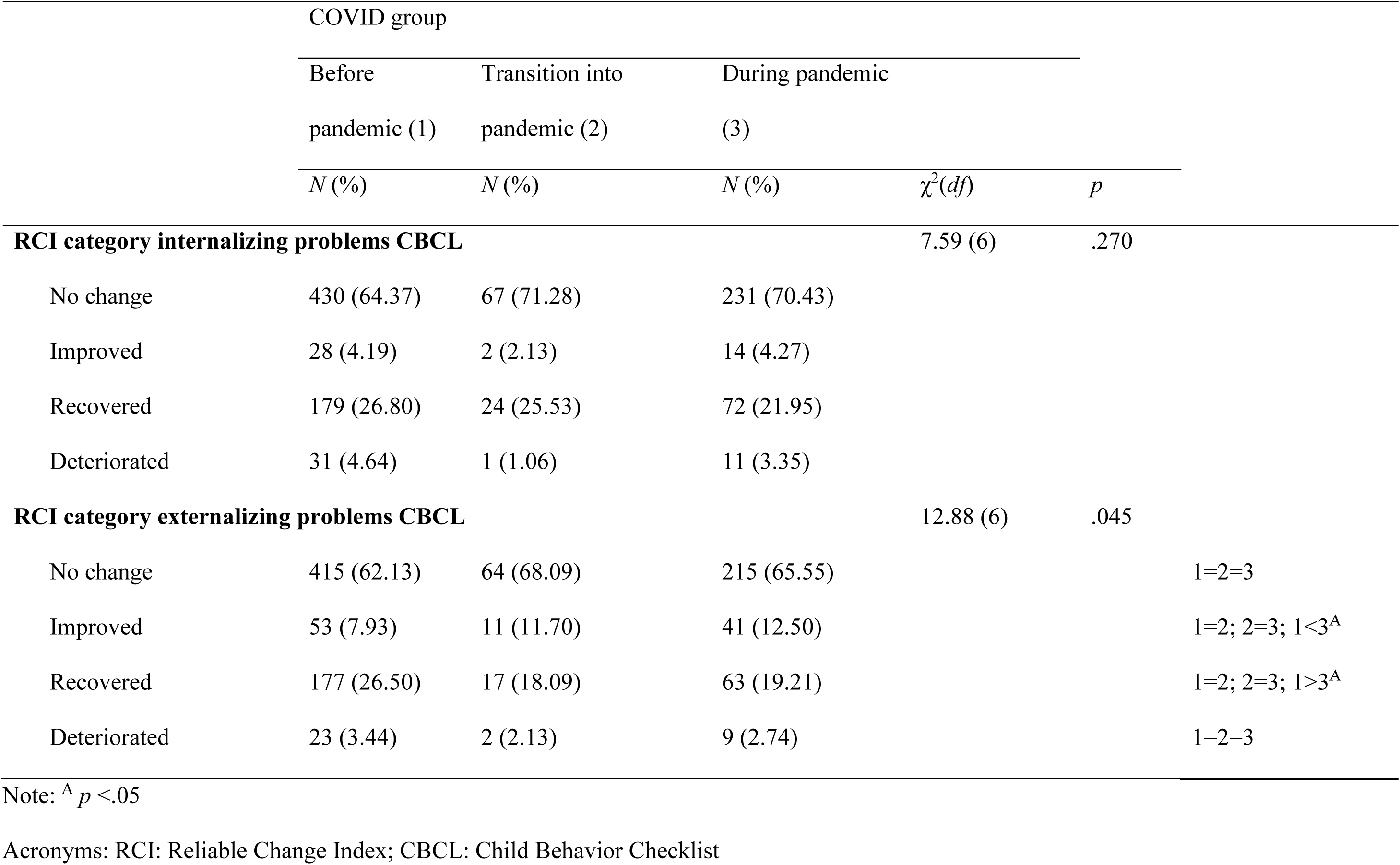
Differences between the three COVID groups in internalizing and externalizing clinical outcome categories based on the Reliable Change Index.

**Table 3.** Differences in internalizing and externalizing problem severity between the three COVID groups at the beginning and end of treatment.

| | COVID group | | | $F(df_{within})^C$ | $p$ | Post-hoc |
| --- | --- | --- | --- | --- | --- | --- |
|  | Before | Transition into | During pandemic |  |  |  |
|  | pandemic (1) | pandemic (2) | (3) |  |  |  |
| | $M (SD)$ | $M (SD)$ | $M (SD)$ | | | |
| Internalizing problems start treatment ($T$ score)<sup>D</sup> | 62.05 (10.26) | 63.51 (8.33) | 64.25 (10.25) | 5.57 (1083) | .004 | 1=2; 1<3<sup>A</sup>; 2=3 |
| Internalizing problems end treatment ($T$ score)<sup>D</sup> | 56.70 (11.47) | 58.06 (10.53) | 59.55 (11.42) | 6.84 (1083) | .001 | 1=2; 1<3<sup>B</sup>; 2=3 |
| Externalizing problems start treatment ($T$ score)<sup>D</sup> | 64.63 (9.93) | 66.89 (9.68) | 66.39 (11.34) | 6.48 (1083) | .002 | 1<2<sup>A</sup>; 1<3<sup>A</sup>; 2=3 |
| Externalizing problems end treatment ($T$ score)<sup>D</sup> | 58.90 (10.94) | 61.54 (10.56) | 61.81 (12.18) | 9.81 (1083) | <.001 | 1<2<sup>A</sup>; 1<3<sup>B</sup>; 2=3 |

Note: <sup>A</sup> $p < .05$ ; <sup>B</sup> $p < .001$ ; <sup>C</sup> $df_{between} = 2$ for all $F$ -tests; <sup>D</sup> Internalizing and externalizing problems at start and end of treatment are reported uncorrected for child's and informant's sex, child's age at start of treatment, and treatment duration.

### Externalizing problems

In all three groups, there was a significant decrease in externalizing problems (*F*(1,1083) = 13.68, *p* < .001, partial η^2^ < .01). The change in externalizing problems from start to end of treatment did not differ between the three groups (*F*(2,1083) = 1.85, *p* = .158, partial η^2^ = .003). See Figure 2B for a graphical representation of these results. Clinical status based on externalizing problems differed between the groups at the end of treatment (χ^2^(6) = 12.88, *p* = .045). Post-hoc analyses showed that fewer children in the “During pandemic” group (19.21%) recovered from externalizing problems than in the “Before pandemic” group (26.50%). However, more children in the “During pandemic” group (12.50%) achieved reliable change than in the “Before pandemic” group (7.93%). No other between-group differences were found to be significant (see Table 2). Externalizing problems at the start (*F*(2,1083) = 6.48, *p* = .002, η^2^ = .01) and at the end (*F*(2,1083) = 8.81, *p* < .001, η^2^ = .02) of treatment were significantly different between the three groups. Post-hoc *t*-tests revealed that problem severity in the “During pandemic” group and in the “Transition into pandemic” group was significantly higher than in the “Before pandemic” group, both at the beginning and end of treatment. No other between-group differences were found to be significant (see Table 3). In addition, externalizing problems at the end of treatment were still at clinical levels for the “During pandemic” and “Transition into pandemic” groups (*T* score > 60; see Figure 2B).

## Discussion

We aimed to investigate if treatment effects and mental health of children receiving youth care in the Netherlands were affected by the COVID-19 pandemic. Mental health was operationalized as internalizing and externalizing problems rated by parents. We used a longitudinal research design in three groups of children that were treated at different time periods relative to the COVID-19 pandemic: “Before pandemic”, “Transition into pandemic” and “During pandemic”. We tested group differences in 1) the reduction in problem scores and changes in clinical status at the end of treatment; 2) reported problem scores when children entered care, and 3) reported problem scores when children left care.

Using a similar approach as de Beurs et al. (2022) who studied a sample of adults, we did not find differences between the three groups of children in the overall reduction in internalizing or externalizing problems over time. All groups did show diminished problems after treatment compared to before treatment. However, we found that fewer children who were treated during the COVID-19 pandemic recovered from externalizing problems than children who were treated prior to the pandemic, although more children in this group achieved reliable change in externalizing problems. These children also still scored at clinical levels of externalizing problems at the end of treatment. These findings show that although treatments seemed to have similar effects throughout the pandemic, fewer children recovered from their externalizing problems compared to before the pandemic.

To further contextualize these findings we extended the design used by de Beurs et al. (2022), by also inspecting the level of problems at the start and end of treatment in the three groups. Here we found that children who entered care during the COVID-19 pandemic experienced more internalizing and externalizing problems than peers who entered care before the pandemic. In addition, we found that children who experienced the transition into the COVID-19 pandemic during their treatment already experienced more externalizing problems when they entered care compared to their peers who were treated entirely before the pandemic. This particular finding cannot be ascribed to the pandemic, as it did not play a role when these children entered youth care. This confirms previous studies showing that child mental health was already declining prior to the COVID-19 pandemic (Bachmann et al., 2016; Collishaw, 2015; Kronström et al., 2023; M. Olfson, Druss, & Marcus, 2015; Patalay & Gage, 2019). Societal changes such as increased social media use (Abi-Jaoude, Naylor, & Pignatiello, 2020; Kelly, Zilanawala, Booker, & Sacker, 2018), increased mental health awareness efforts (Foulkes & Andrews, 2023), increasing rates of parental mental health problems (Mark Olfson, Blanco, Wang, Laje, & Correll, 2014; Thapar, Collishaw, Pine, & Thapar, 2012), increased long-term poverty (Chaudry & Wimer, 2016; Ridley, Rao, Schilbach, & Patel, 2020), increased school pressure (Steare, Gutiérrez Muñoz, Sullivan, & Lewis, 2023), and earlier puberty onset (Golub et al., 2008; Lee & Styne, 2013) have previously been linked to these trends of poorer child mental health over the past decades. The increased internalizing and externalizing problems in children who entered care during the pandemic in our sample may therefore be a continuation of changes already occurring at a larger societal scale, rather than only being a result of the pandemic on child mental health.

Furthermore, we found that children who entered care during the COVID-19 pandemic ended treatment with higher internalizing and externalizing problems than their peers who were treated before the pandemic. In addition, children who experienced the transition into the pandemic also showed higher externalizing problems at the end of their treatment compared to peers treated before the pandemic. This is in line with previous findings that showed poorer child mental health during the pandemic compared to before the pandemic (Adegboye et al., 2021; Cost et al., 2022; Crandal et al., 2022; Gilsbach, Herpertz-Dahlmann, & Konrad, 2021; Lopez-Serrano et al., 2021; J. Zijlmans et al., 2021; Josjan Zijlmans et al., 2023) and is potentially partially due to true effects of the pandemic on youth care. A substantial portion of child mental health care had to transition into telehealth early in the pandemic due to social distancing measures (Folk et al., 2022; Palinkas et al., 2021; Revet et al., 2023). Yet, organizations often encountered difficulties early in this process with licensing, infrastructure, or availability of devices for both clinicians and families receiving care (Bierbooms et al., 2020; Feijt et al., 2020; Folk et al., 2022; Palinkas et al., 2021; Revet et al., 2023), which may have disrupted treatment. We did find a substantially longer treatment duration in the transitional group, suggesting that this group needed more time to achieve the same therapeutic effects as children treated before or entirely during the pandemic. This may indeed be due to difficulties adjusting to COVID-19 restrictions in treatment. However, as we have limited background information regarding possible changes to treatment it is also a possibility that treatments were more spread out, and that children in the transitional group received the same amount of care in a longer period of time as children treated before and entirely during the pandemic. More long-term detrimental effects of COVID-19 on the mental healthcare sector have also been reported, such as persisting increased psychological problems (Alexiou et al., 2021; Crocker et al., 2023; van Doesum et al., 2023) and poorer working conditions (Alexiou et al., 2021; Crocker et al., 2023; Palinkas et al., 2021; Revet et al., 2023; van Doesum et al., 2023) among mental healthcare workers due to the pandemic.

Although all three groups achieved similar reductions in internalizing and externalizing problems over time, our findings do suggest that children treated during the pandemic left care with more problems than before the pandemic. Children who were treated either partially or entirely during the pandemic on average still score at clinical levels of externalizing problems at the end of treatment, whereas this was not the case in our pre-pandemic sample. Although it is unclear whether this is due to the COVID-19 pandemic alone, or is also due to pre-existing societal changes in child mental health, these findings do suggest that using a different approach in treating these children may be beneficial, as their problems before entering treatment are more severe compared to before the pandemic.

### Strengths and limitations

This is the first longitudinal study in which the effects of the COVID-19 pandemic on youth care were investigated using repeated measures obtained in both a group of children who were treated entirely before the COVID-19 pandemic and children treated during the pandemic. Previous prospective studies that included pre-pandemic measurements often did not include children treated entirely during or entirely before the pandemic, limiting the conclusions that could be drawn from their findings. In addition, many studies in which findings from before and during the pandemic were compared were cross-sectional in nature, and could therefore not examine within-person effects of the pandemic on children. We were able to address these limitations by using both repeated measures and three groups of children treated at different time periods relative to the COVID-19 pandemic. In addition, by including children who experienced the transition into COVID-19 measures, we were also able to further understand how the pandemic affected child mental health and treatment (e.g., treatment duration) independently of changes to child mental health that were already occurring before the pandemic. We could also include large samples of children and employed reliable and valid instruments to measure child mental health.

Although our study included a large sample that covered an extensive period, we had limited background information about our sample. Besides treatment duration no information was available regarding adjustments to treatment, such as delays to treatment or how much care transitioned to telehealth. In addition, we had limited demographic information available to assess generalizability or to control for any confounding influences. Conclusions about the generalizability of our sample are further limited as the CBCL is not filled in for every child that enters the youth care institutions that are part of the Learning Database Youth. Selection effects are therefore a possibility.

### Future directions

Although we covered the entire duration of the pandemic, the possible long-term effects of the pandemic cannot be gauged based on our study. Future studies should compare children who entered care after the COVID-19 pandemic (i.e. after restriction measures ended) to children treated before and during the pandemic, to study whether children’s mental health problems are returning to pre-pandemic levels. In addition, although our study showed that many children are leaving care with clinical levels of problems, it is unclear how these children are doing now. Future studies might study how these groups of children are currently coping.

In addition, to further understand the effects of the COVID-19 pandemic on treatments, further studies might be conducted regarding specific adjustments to care made as a direct result of pandemic restrictions. Future studies might investigate the influence of factors such as transitions to telehealth and poorer mental health and working conditions of mental healthcare workers on treatment effects. In addition, factors such as socioeconomic status, treatment dropout, and specific types of mental health problems might be evaluated. This information is necessary to better understand the potential effects of these factors on the mental healthcare and problems of youth during global health-related disasters such as pandemics. As previous research has shown that poor child mental health may lead to lifelong negative consequences such as overall poor health and negative social and economic outcomes (Goodman, Joyce, & Smith, 2011), it is important to further understand how child mental health might be affected by such disrupting global events, both during these events and afterwards.

### Conclusions

To conclude, we did not find indications that treatment effects were affected by the COVID-19 pandemic. However, we did find elevated internalizing and externalizing problem severity in children treated entirely during the pandemic, and elevated externalizing problem severity in children who started treatment during the pandemic compared to children treated before the pandemic. Our findings indicate that child mental health has deteriorated since the start of the pandemic, possibly as a result of both pre-pandemic trends of deteriorating child mental health and due to the COVID-19 pandemic specifically. As poor mental health at a young age may predict lifelong negative consequences, it is important to study the long-term development and treatment of child mental health.

## KEY POINTS AND RELEVANCE

*N = 97 words*

- The COVID-19 pandemic severely disrupted children’s daily lives, but also treatment provision and delivery for children receiving mental healthcare. Mental health problems of children receiving such care has deteriorated since the pandemic.
- Our findings show that the pandemic has not affected treatment effectiveness. However, children’s mental health problems have increased, and children are leaving care with more problems than before the pandemic.
- As children seem to enter and leave care with more problems compared to before the pandemic, our findings beg the question whether current treatment provision and delivery is sufficient to deal with these heavier burdens.

## Data Availability

All data produced in the present study are available upon reasonable request to the authors

## Abbreviations

CBCL: Child Behavior Checklist
RCI: Reliable Change Index
COVID-19: coronavirus disease 2019

## ACKNOWLEDGEMENTS

We thank all participating families. This research was funded by ZonMw project number 50-56300-98-973. We thank all organizations participating in the Partnership for Effective Youth Care in the Netherlands (SEJN) for sharing their data through the Learning Database Youth. We also thank the COVID-19 Wellbeing and Mental Health Consortium for their valuable contribution to this study.

## References

Abi-Jaoude, E., Naylor, K. T., & Pignatiello, A. (2020). Smartphones, social media use and youth mental health. Canadian Medical Association Journal, 192(6), E136–E141.

Achenbach, T. M., & Rescorla, L. A. (2001). Child behavior checklist for ages 6-18: University of Vermont Burlington, VT.

Achenbach, T. M., & Rescorla, L. A. (2007). Multicultural supplement to the manual for the ASEBA school-age forms & profiles. Burlington VT: University of Vermont Research Center for Children, Youth, & Families.

Adegboye, D., Williams, F., Collishaw, S., Shelton, K., Langley, K., Hobson, C., et al. (2021). Understanding why the COVID-19 pandemic-related lockdown increases mental health difficulties in vulnerable young children. JCPP Advances, 1(1), e12005.

Alexiou, E., Steingrimsson, S., Akerstrom, M., Jonsdottir, I. H., Ahlstrom, L., Finizia, C., et al. (2021). A survey of psychiatric healthcare workers’ perception of working environment and possibility to recover before and after the first wave of COVID-19 in Sweden. Frontiers in Psychiatry, 12, 770955.

Bachmann, C. J., Aagaard, L., Burcu, M., Glaeske, G., Kalverdijk, L. J., Petersen, I., et al. (2016). Trends and patterns of antidepressant use in children and adolescents from five western countries, 2005-2012. European Neuropsychopharmacology 26(3), 411–419.

Bierbooms, J., van Haaren, M., W.A., I. J., de Kort, Y. A. W., Feijt, M., & Bongers, I. M. B. (2020). Integration of online treatment into the “new normal” in mental health care in post-COVID-19 times: Exploratory qualitative Study. JMIR Formative Research, 4(10), e21344.

Branquinho, C., Santos, A. C., Noronha, C., Ramiro, L., & de Matos, M. G. (2022). COVID-19 pandemic and the second lockdown: The 3rd wave of the disease through the voice of youth. Child Indicators Research, 15(1), 199–216.

Bruinen de Bruin, Y., Lequarre, A.-S., McCourt, J., Clevestig, P., Pigazzani, F., Zare Jeddi, M., et al. (2020). Initial impacts of global risk mitigation measures taken during the combatting of the COVID-19 pandemic. Safety Science, 128, 104773.

Caroppo, E., Mazza, M., Sannella, A., Marano, G., Avallone, C., Claro, A. E., et al. (2021). Will nothing be the same again? Changes in lifestyle during COVID-19 pandemic and consequences on mental health. International Journal of Environmental Research and Public Health, 18(16), 8433.

Chaudry, A., & Wimer, C. (2016). Poverty is not just an indicator: The relationship between income, poverty, and child well-being. Academic Pediatrics, 16(3), S23–S29.

Collishaw, S. (2015). Annual research review: Secular trends in child and adolescent mental health. Journal of Child Psychology and Psychiatry 56(3), 370–393.

Cost, K. T., Crosbie, J., Anagnostou, E., Birken, C. S., Charach, A., Monga, S., et al. (2022). Mostly worse, occasionally better: impact of COVID-19 pandemic on the mental health of Canadian children and adolescents. European Child & Adolescent Psychiatry, 31(4), 671–684.

Crandal, B. R., Hazen, A. L., Dickson, K. S., Tsai, C. K., Trask, E. V., & Aarons, G. A. (2022). Mental health symptoms of youth initiating psychiatric care at different phases of the COVID-19 pandemic. Child and Adolescent Psychiatry and Mental Health, 16(1), 77.

Crocker, K. M., Gnatt, I., Haywood, D., Butterfield, I., Bhat, R., Lalitha, A. R. N., et al. (2023). The impact of COVID-19 on the mental health workforce: A rapid review. International Journal of Mental Health Nursing, 32(2), 420–445.

de Beurs, E., Blankers, M., Peen, J., Rademacher, C., Podgorski, A., & Dekker, J. (2022). Impact of COVID-19 social distancing measures on routine mental health care provision and treatment outcome for common mental disorders in the Netherlands. Clinical Psychology & Psychotherapy, 29(4), 1342–1354.

Feijt, M., de Kort, Y., Bongers, I., Bierbooms, J., Westerink, J., & W, I. J. (2020). Mental health care goes online: Practitioners’ experiences of providing mental health care during the COVID-19 pandemic. Cyberpsychology, Behavior, and Social Networking, 23(12), 860–864.

Folk, J. B., Schiel, M. A., Oblath, R., Feuer, V., Sharma, A., Khan, S., et al. (2022). The transition of academic mental health clinics to telehealth during the COVID-19 pandemic. Journal of the American Academy of Child & Adolescent Psychiatry, 61(2), 277–290.

Foulkes, L., & Andrews, J. L. (2023). Are mental health awareness efforts contributing to the rise in reported mental health problems? A call to test the prevalence inflation hypothesis. New Ideas in Psychology, 69, 101010.

Gilsbach, S., Herpertz-Dahlmann, B., & Konrad, K. (2021). Psychological impact of the COVID-19 pandemic on children and adolescents with and without mental disorders. Frontiers in Public Health, 9, 679041.

Golub, M. S., Collman, G. W., Foster, P. M. D., Kimmel, C. A., Rajpert-De Meyts, E., Reiter, E. O., et al. (2008). Public health implications of altered puberty timing. Pediatrics, 121(3), S218–S230.

Goodman, A., Joyce, R., & Smith, J. P. (2011). The long shadow cast by childhood physical and mental problems on adult life. Proceedings of the National Academy of Sciences, 108(15), 6032–6037.

Jacobson, N. S., & Truax, P. (1991). Clinical significance: A statistical approach to defining meaningful change in psychotherapy research. Journal of Consulting and Clinical Psychology, 59(1), 12–19.

Kauhanen, L., Wan Mohd Yunus, W. M. A., Lempinen, L., Peltonen, K., Gyllenberg, D., Mishina, K., et al. (2022). A systematic review of the mental health changes of children and young people before and during the COVID-19 pandemic. European Child & Adolescent Psychiatry, 32(6), 995–1013.

Kelly, Y., Zilanawala, A., Booker, C., & Sacker, A. (2018). Social media use and adolescent mental health: Findings from the UK Millennium Cohort Study. EClinicalMedicine, 6, E59–E68.

Khanna, R. C., Cicinelli, M. V., Gilbert, S. S., Honavar, S. G., & Murthy, G. S. V. (2020). COVID-19 pandemic: Lessons learned and future directions. Indian Journal of Ophthalmology, 68(5), 703–710.

Kronström, K., Tiiri, E., Vuori, M., Ellilä, H., Kaljonen, A., & Sourander, A. (2023). Multi-center nationwide study on pediatric psychiatric inpatients 2000–2018: length of stay, recurrent hospitalization, functioning level, suicidality, violence and diagnostic profiles. European Child & Adolescent Psychiatry, 32(5), 835–846.

Lee, Y., & Styne, D. (2013). Influences on the onset and tempo of puberty in human beings and implications for adolescent psychological development. Hormones and Behavior, 64(2), 250–261.

Lopez-Serrano, J., Diaz-Boveda, R., Gonzalez-Vallespi, L., Santamarina-Perez, P., Bretones-Rodriguez, A., Calvo, R., et al. (2021). Psychological impact during COVID-19 lockdown in children and adolescents with previous mental health disorders. Revista de Psiquiatría y Salud Mental (English Edition), 16(1), 32–41.

Olfson, M., Blanco, C., Wang, S., Laje, G., & Correll, C. U. (2014). National trends in the mental health care of children, adolescents, and adults by office-based physicians. JAMA psychiatry, 71(1), 81–90.

Olfson, M., Druss, B. G., & Marcus, S. C. (2015). Trends in mental health care among children and adolescents. The New England Journal of Medicine, 372(21), 2029–2038.

Palinkas, L. A., De Leon, J., Salinas, E., Chu, S., Hunter, K., Marshall, T. M., et al. (2021). Impact of the COVID-19 Pandemic on Child and Adolescent Mental Health Policy and Practice Implementation. International Journal of Environmental Research and Public Health, 18(18), 9622.

Panchal, U., Salazar de Pablo, G., Franco, M., Moreno, C., Parellada, M., Arango, C., et al. (2021). The impact of COVID-19 lockdown on child and adolescent mental health: systematic review. European Child & Adolescent Psychiatry, 32(7).

Patalay, P., & Gage, S. H. (2019). Changes in millennial adolescent mental health and health-related behaviours over 10 years: a population cohort comparison study. International Journal of Epidemiology, 48(5), 1650–1664.

Revet, A., Hebebrand, J., Anagnostopoulos, D., Kehoe, L. A., Gradl-Dietsch, G., Covid-Child Adolescent Psychiatry, C., et al. (2023). Perceived impact of the COVID-19 pandemic on child and adolescent psychiatric services after 1 year (February/March 2021): ESCAP CovCAP survey. European Child & Adolescent Psychiatry 32(2), 249–256.

Ridley, M., Rao, G., Schilbach, F., & Patel, V. (2020). Poverty, depression, and anxiety: Causal evidence and mechanisms. Science, 370(6522), eaay0214.

RIVM. (2023a). Tijdlijn van coronamaatregelen 2020. Retrieved 10 July, 2023, from https://www.rivm.nl/gedragsonderzoek/tijdlijn-van-coronamaatregelen-2020

RIVM. (2023b). Tijdlijn van coronamaatregelen 2021. Retrieved 10 July, 2023, from https://www.rivm.nl/gedragsonderzoek/tijdlijn-van-coronamaatregelen-2021

RIVM. (2023c). Tijdlijn van coronamaatregelen 2022. Retrieved 10 July, 2023, from https://www.rivm.nl/gedragsonderzoek/tijdlijn-maatregelen-covid-2022

Samji, H., Wu, J., Ladak, A., Vossen, C., Stewart, E., Dove, N., et al. (2022). Review: Mental health impacts of the COVID-19 pandemic on children and youth – a systematic review. Child and Adolescent Mental Health, 27(2), 173–189.

SEJN. (2020). De Lerende Databank Jeugd - Betere benutting van gegevens door gebruik van een feedbacksysteem [The Learning Database Youth: Better utilisation of data by use of a feedback system].

Singh, S., Roy, D., Sinha, K., Parveen, S., Sharma, G., & Joshi, G. (2020). Impact of COVID-19 and lockdown on mental health of children and adolescents: A narrative review with recommendations. Psychiatry Research, 293, 113429.

Steare, T., Gutiérrez Muñoz, C., Sullivan, A., & Lewis, G. (2023). The association between academic pressure and adolescent mental health problems: A systematic review. Journal of Affective Disorders, 339, 302–317.

Thapar, A., Collishaw, S., Pine, D. S., & Thapar, A. K. (2012). Depression in adolescence. The Lancet, 379(9820), 1056–1067.

van Doesum, T. J., Shields-Zeeman, L. S., Leone, S. S., van Meijel, B., Jabbarian, L. J., & van Bon-Martens, M. (2023). Impact of the COVID-19 pandemic on working conditions and mental well-being of mental health professionals in the Netherlands: a cross-sectional study. BMJ Open, 13(4), e062242.

Veerman, J. W., & De Meyer, R. E. (2015). Consistency of outcomes of home-based family treatment in The Netherlands as an indicator of effectiveness. Children and Youth Services Review, 59, 113–119.

Zijlmans, J., Teela, L., van Ewijk, H., Klip, H., van der Mheen, M., Ruisch, H., et al. (2021). Mental and social health of children and adolescents with pre-existing mental or somatic problems during the COVID-19 pandemic lockdown. Frontiers in Psychiatry, 12, 692853.

Zijlmans, J., Tieskens, J. M., van Oers, H. A., Alrouh, H., Luijten, M. A., de Groot, R., et al. (2023). The effects of COVID-19 on child mental health: Biannual assessments up to April 2022 in a clinical and two general population samples. JCPP Advances, 3(2), e12150.

