## Supplemental Table 1 for "Changes in internalizing and externalizing problems in Dutch children and adolescents receiving youth care before and during the COVID-19 pandemic"

**Supplementary materials**

**Table 1**

*Correlations between study (internalizing and externalizing problems before and after treatment) and control (child's age at start of treatment, treatment duration, sex of both the child and informant) variables*

| | 1 | 2 | 3 | 4 | 5 | 6 | 7 | 8 |
|---|---|---|---|---|---|---|---|---|
| 1. Internalizing problems before treatment (*T* score) | - | | | | | | | |
| 2. Internalizing problems after treatment (*T* score) | .64$^{B}$ | - | | | | | | |
| 3. Externalizing problems before treatment (*T* score) | .49$^{B}$ | .37$^{B}$ | - | | | | | |
| 4. Externalizing problems after treatment (*T* score) | .36$^{B}$ | .61$^{B}$ | .71$^{B}$ | - | | | | |
| 5. Age at start of treatment | .07$^{A}$ | .11$^{B}$ | .10$^{B}$ | .13$^{B}$ | - | | | |
| 6. Treatment duration (days) | .02 | -.02 | -.08$^{B}$ | -.08$^{B}$ | -.27$^{B}$ | - | | |
| 7. Sex of child | .10$^{B}$ | .04 | -.05 | -.05 | -.002 | 0.02 | - | |

| | | | | | | | | |
|---|---|---|---|---|---|---|---|---|
| 8. Sex of informant | .09[B] | .05 | .12[B] | .06[A] | -.01 | -.01 | -.02 | - |

Note: [A] $p$ <.05; [B] $p$ <.001
